# A web tool for the exploration of hospital prescription data in England

**DOI:** 10.1101/2023.12.14.23299817

**Authors:** Theo Sanderson

## Abstract

Medicine prescription data is a valuable resource, with the power to reveal insights into healthcare trends, cost efficiencies, and geographic disparities. In England, primary care prescription data has been openly accessible for analysis for some time through a web tool, providing significant benefits. Since 2020, the National Health Service in England has also released data on secondary care prescriptions, i.e. prescribing within hospitals. This is an important dataset, but until now has been available only in a raw form that requires considerable technical skills to be used for even the analysis of basic trends. I have built a web application that enables anyone to easily analyse trends in this data, which is available at hospitalprescriptions.genomium.org.

## Introduction

Prescription data provides an important reservoir of data for potential insights into drug utilisation patterns, healthcare practices, and public health trends. Since 2015, the Open Prescribing project has provided a comprehensive platform for the analysis of primary care prescription data in England (OpenPrescribing.net, Curtis and Goldacre (2018)). It now receives >130,000 unique visitors per year, and has made substantial impacts on clinical practice (Walker et al., 2019).

Historically, secondary care prescription data – i.e. data on prescriptions in hospitals – had not been made publicly available. The Bennett Institute, which builds OpenPrescribing, called in 2020 for secondary care data (which had for some time been collected by a company called Rx-info), to be made publicly available (Goldacre and MacKenna, 2020). In parallel (as described in the online responses to that article), an arrangement was developed for Rx-info’s dataset to be made available through the NHSBSA’s Open Data Portal as the Secondary Care Medicines Data (SCMD) dataset, which was launched soon after in the same year.

The SCMD has been an important dataset, and has been used, for example as a denominator to estimate the rate of adverse drug reports (Sandhu et al., 2021), and to investigate asthma prescribing (Rowan and MacKenna, 2020). It has widespread potential applications: my own interest in this area came from my attempts to link temporal and geographic trends (primarily at the level of countries), between a mutagenic drug and the rate of specific mutational events in virus genomes (Sanderson et al., 2023).

However, the SCMD dataset requires significant processing in order to extract useful insights. For example it is available as a set of CSV files, one per month, which must each be downloaded and then combined in order to generate temporal insights. The combined dataset currently comprises 17 million rows of data. Many analyses require combining the dataset with other information – hospitals are indicated by their ODS code, e.g. *R1H*, but users are likely to want to translate this to its textual value, e.g. *Barts Health NHS Trust*. Similarly any aggregation by drug *ingredient* (e.g. *aspirin*), instead of specific formulation (e.g. *aspirin 500mg granules sachets sugar free*) requires joining to the NHS Dictionary of Medicines and Devices (dm+d).

Here, I describe a web tool for exploration of this dataset. This tool permits users to inspect temporal trends in hospital prescriptions, both nationally and at the level of individual NHS Trusts, and permits aggregation by ingredient. It is available at hospitalprescriptions.genomium.org.

## Application features

The web application has two main modes. The *Formulation* mode allows users to search for specific “Virtual Medicine Products” which are genericised versions of real products, for example ‘Ibuprofen 200mg capsules’. When the user selects a product, a graph of its national trends over time is shown, which can be visualised as a bar plot or as a line plot (with or without smoothing). Users can choose to view either the number of items prescribed (i.e. tablets, vials, etc.) or their ‘indicative cost’ – though the latter metric does not represent the real cost paid and sometimes appears to be affected by data entry errors, so may not be especially useful.

The *Ingredient* mode allows selection of any ingredient of medicine products (i.e. typically a drug, e.g. ‘Ibuprofen’) - it then aggregates the total usage of this ingredient, generally in grams, across all products. This means that if a hospital switches from prescribing 2,000 50 mg tablets per month to prescribing 1,000 100 mg tablets, the resulting graph reflects that the total usage of the drug is unchanged.

In *Formulation* mode, users can break down usage into different hospitals, both by presenting the national picture coloured by hospital trust, or by filtering to an individual trust. This is also possible in the *Ingredient* mode, which adds additional features to break down usage by the specific product in which the ingredient is found, and by the route of administration of the product (i.e. oral, intravenous, etc.)

### Implementation

I created a Postgres database instance to house SCMD data (the final size of the table is >1.3 GB) and related datasets from the dm+d and about hospitals. I populate it using a script which I make openly available. It performs the following operations:

- downloads each month’s data from the SCMD dataset
- downloads the NHS dm+d: this requires a manual login to TRUD, although it should be automatable (OpenPrescribing code)
- downloads hospital trust ODS mappings, giving the names of hospital trusts from their ODS code
- creates a series of database tables using the above data
- adds a table representing common units, and their mappings to standard units. (e.g. *mg* are mapped to *0*.*001 g*)
- Adds a series of indexes to the database to speed up queries

The web application is implemented in NextJS. It uses backend API routes that query the Postgres database, and a frontend implemented in React. The most complex database queries are for ingredient-based searches, with database joins used to connect a specific ingredient to all of the specific formulations in which it appears, with its strength in each of the formulation accounted for and then aggregated by month and by hospital.

The frontend is React-based. Graphing is carried out with Observable Plot (ObservableHQ, 2023). Features for exporting data and downloading an SVG from the graph are available. State is captured in the URL to permit specific graphs to be bookmarked.

### Examples

In this section I will briefly discuss some trends visible in this dataset, to give some sense of its potential utility. In each case I provide a graph from the web application (though graphs are much better viewed live, with tooltips providing additional data).

For a quick sense-check on the dataset and its processing, I analysed drugs we expect to have temporal variation. For example, the anti-influenza drug oseltamivir (Tamiflu) shows winter spikes, apart from during the SARS-CoV-2 pandemic, in which social distancing largely suppressed influenza transmission.

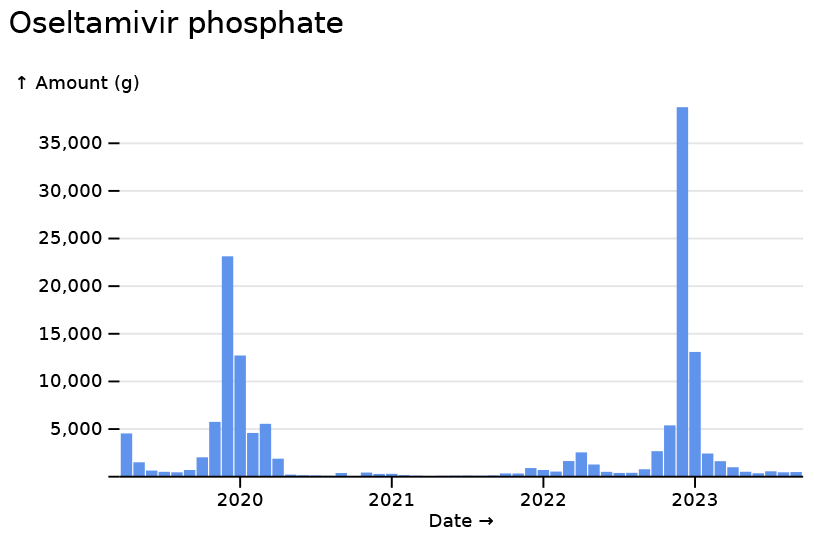

Similarly, palivizumab a monoclonal antibody used pro-phylactically to protect again RSV, but only during its transmission season, shows its expected pattern.

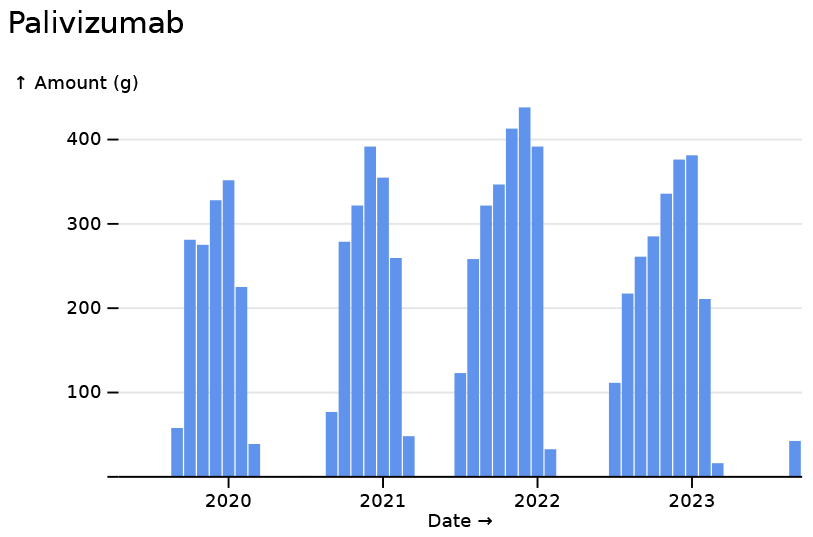

We can also plot the usage of a drug where we expect less variation, such as paracetamol (acetaminophen), with a relatively constant ∼10 tons used per month, with a moderate dent made by the COVID pandemic.

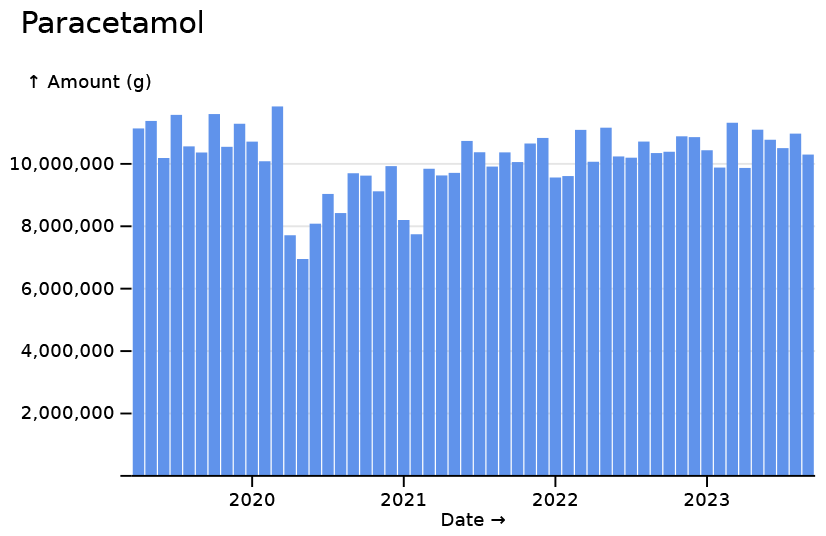

In contrast, ibuprofen shows a more marked reduction during the first two major SARS-CoV-2 waves (and also usage that never fully returns to prepandemic levels).

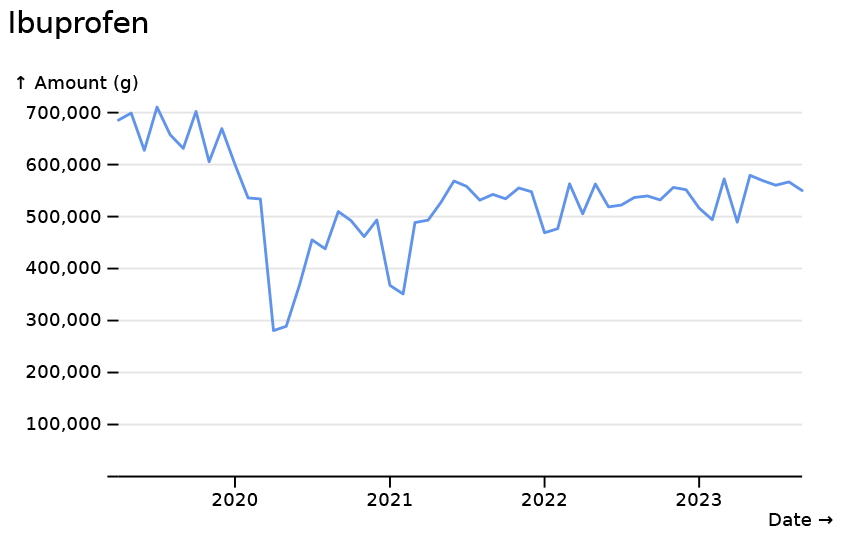

The first two waves of SARS-CoV-2 are highly visible in the dataset. Propofol, used as an anaesthetic for patients receiving mechanical ventilation, sees marked increases during early SARS-CoV-2 waves.

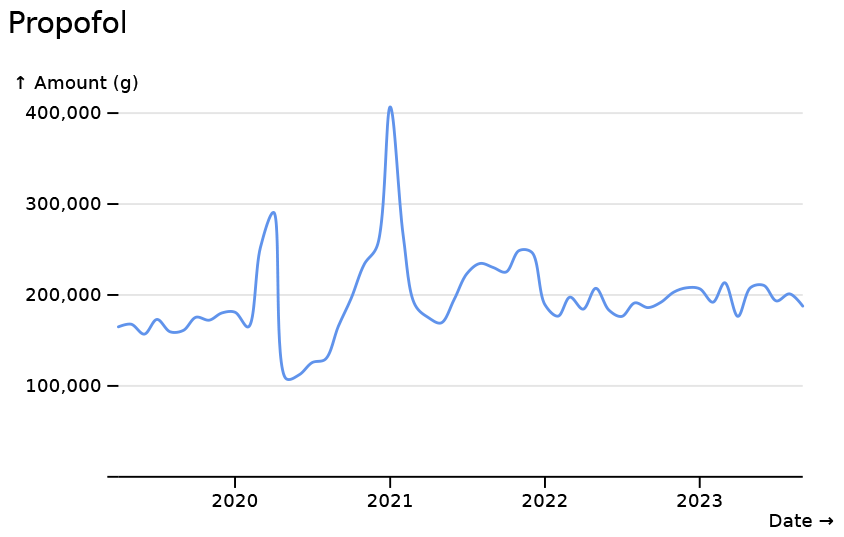

For dexamethasone we see a decrease from baseline during the first COVID-19 wave, and then a marked increase during the second wave, following the discoveries of the RECOVERY trial (The RECOVERY Collaborative Group, 2021).

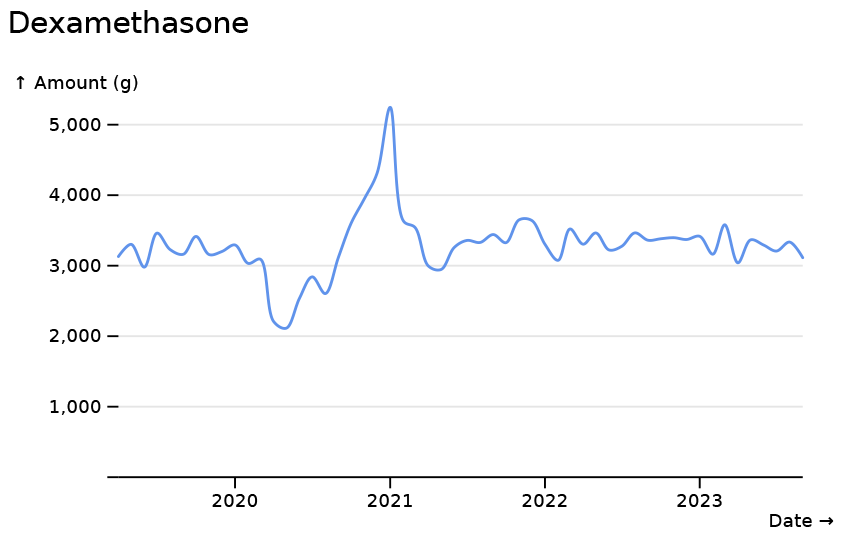

While some kinds of healthcare increased during the pandemic, others decreased. A profound reduction in the use of basiliximab, a monoclonal antibody used immediately following organ transplants to prevent rejection, can be seen during initial SARS-CoV-2 waves. The graph below is coloured by NHS Trust.

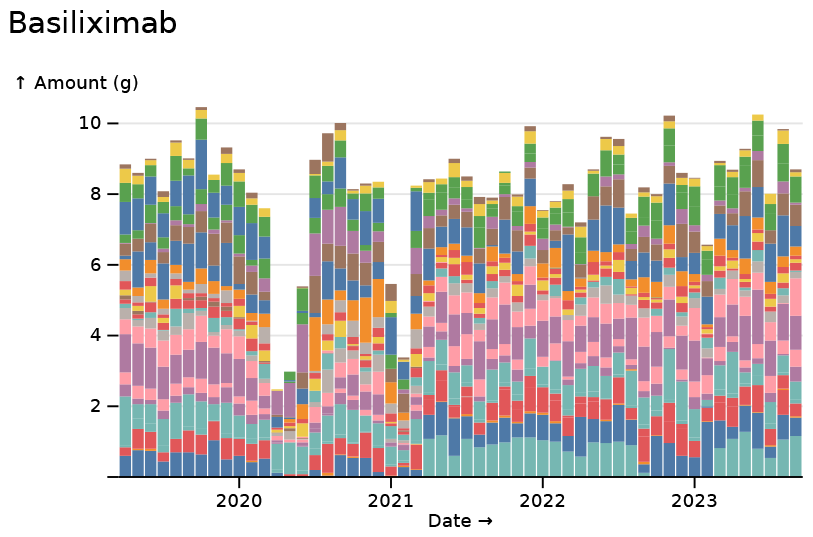

Some reductions are encouraging. Below we can see reductions in the use of anaesthetic gas desflurane. Short-term reductions due to SARS-CoV-2 are visible, but the much larger trend of reduction reflects phasing out due to desflurane’s potential to contribute to global warming: by some measures it is 3,714 times as potent as carbon dioxide in its contribution (Ryan and Nielsen, 2010).

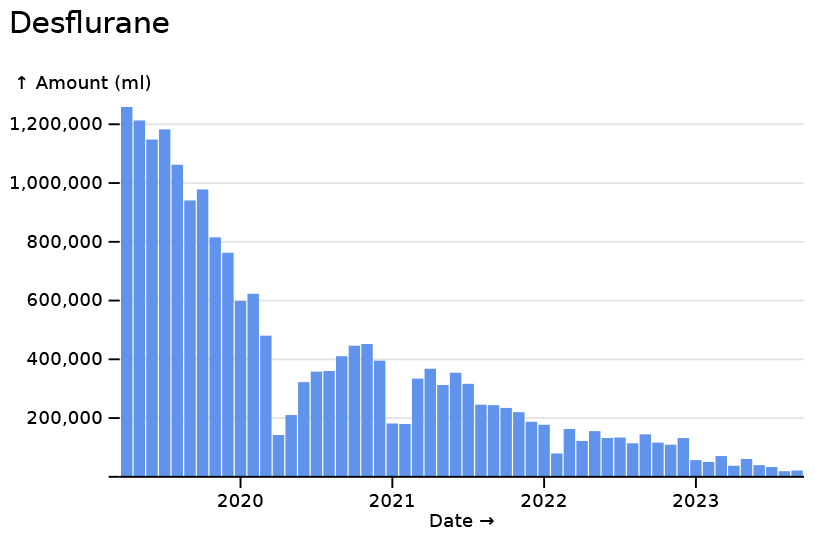

The ability to break down data by the route of drug administration can be important. Waves of SARS-CoV-2 saw a reduction in orally administered morphine (yellow), but increases in intravenous administration (red).

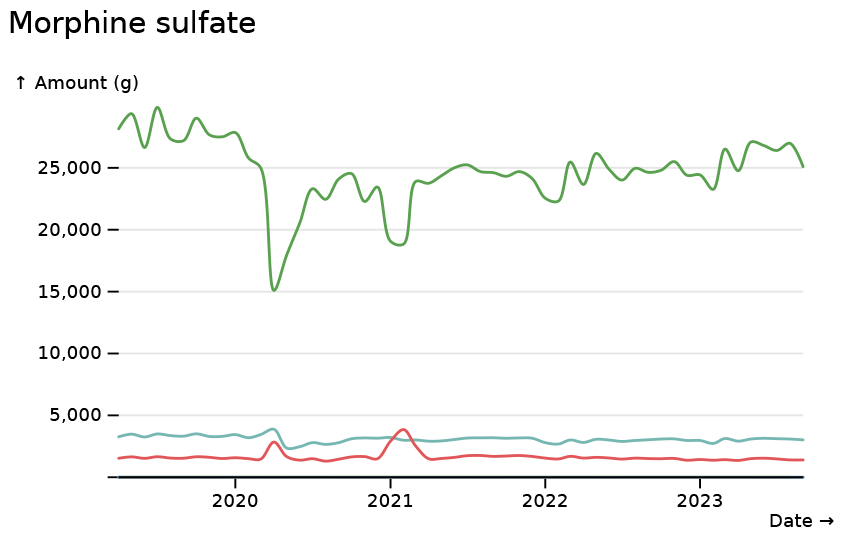

Antibiotic trends show temporal trends, with an increase in the amount of piperacillin prescribed.

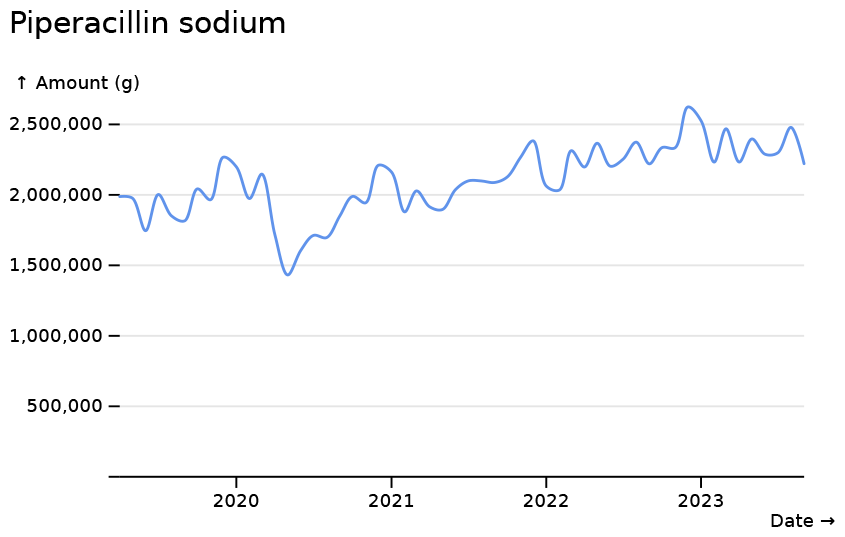

Amoxicillin shows clear increases, across trusts, during the streptococcus group A outbreak of winter 2022-2023 (UK Health Security Agency, 2023).

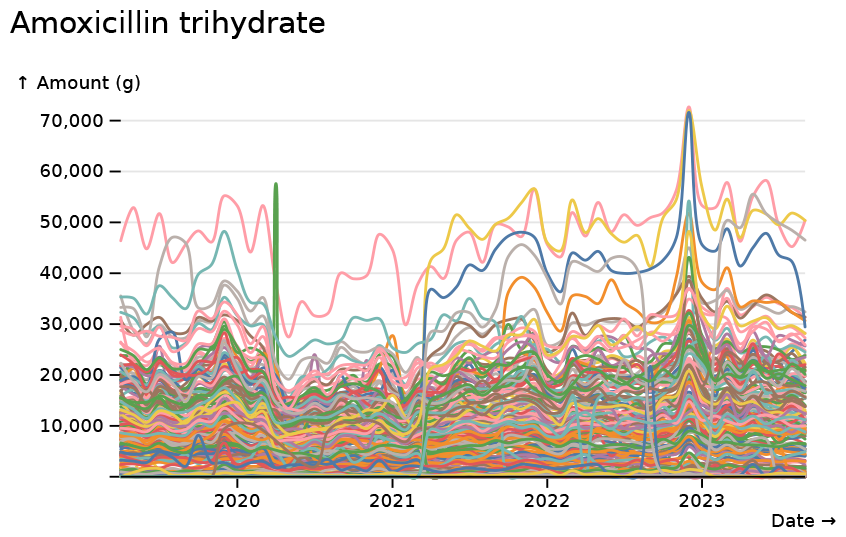

Breaking this down by formulation reveals a particular striking trend for 250mg doses, which reflect prescriptions to young children.

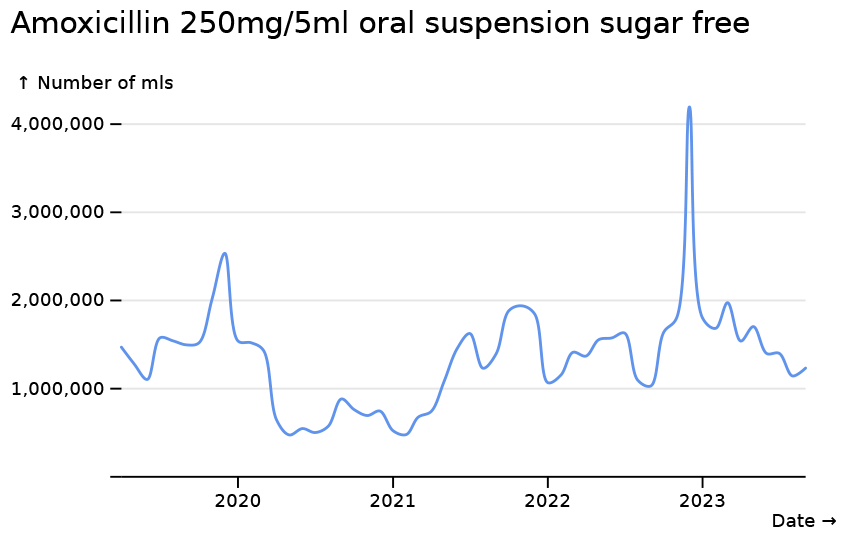

Many medicines show increases during the time period for which data is available. Following a decline in prescribing of methylphenidate hydrochloride, a drug for the treatment of ADHD, at the start of the pandemic, prescriptions have since increased substantially.

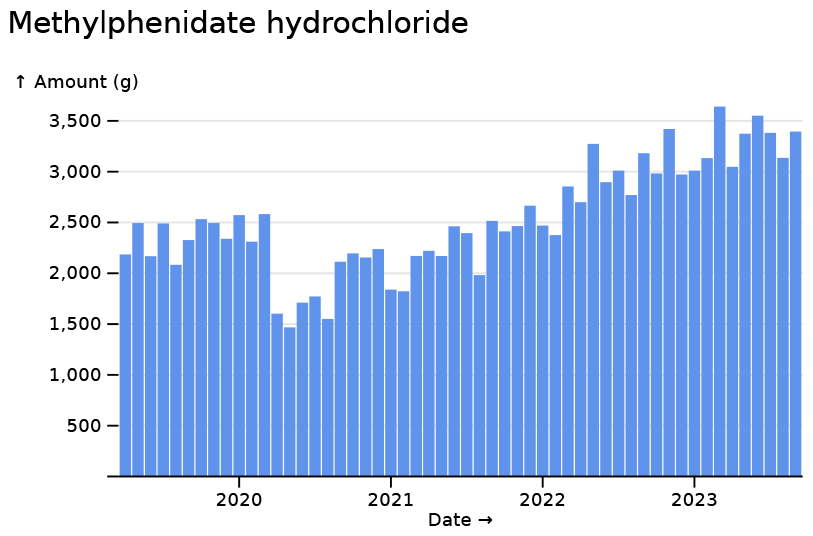

A range of recently developed or licensed drugs show increases reflecting a national roll-out. A whole host of monoclonal antibodies (a text search for “mab” in Ingredients mode is one quick shortcut) show great increases during the period.

Breaking down erenumab (a migraine medication) prescriptions by trust shows an initial period where prescriptions were predominantly at St Thomas’ hospital (in yellow), and then a wide rollout following a NICE recommendation.

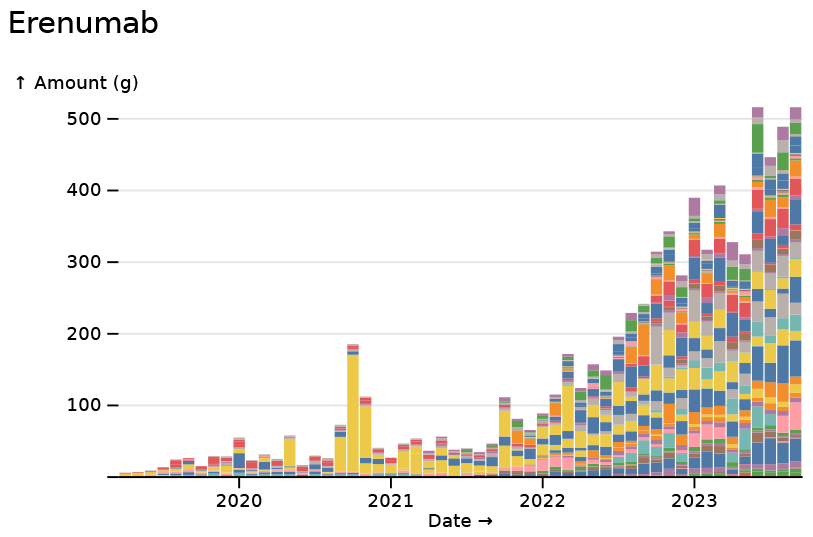

The novel anti-diabetes drug semaglutide begins use during the covered period (but also recent shortages).

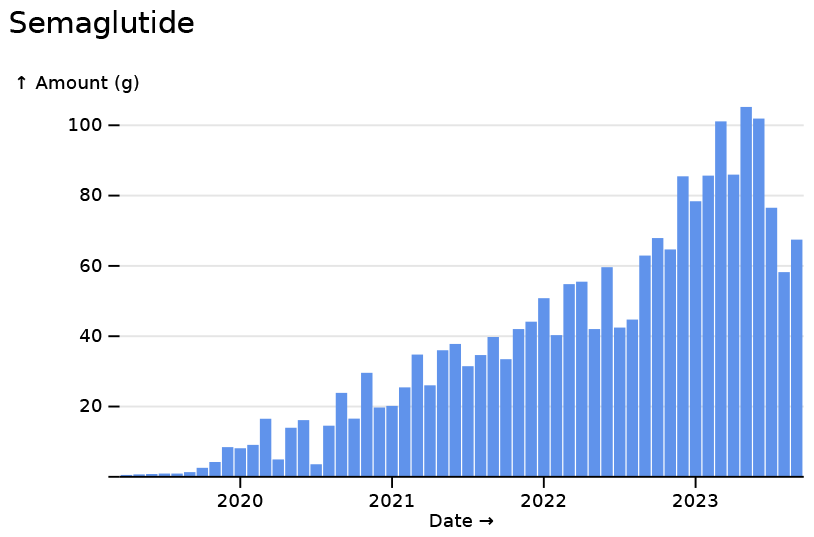

## Limitations

My tool has many limitations. It does not provide normalisation to patient numbers, so is limited in its potential to allow comparisons across hospitals. It does not normalise for standard dosages and does not allow graph with multiple drugs, limiting the ability to compare drugs. It also does not allow grouping drugs into classes. It does not directly display geographic trends. There are also limitations of the underlying dataset. The ‘indicative cost’ metric does not reflect the actual amount paid, due to confidential discounts. There also appear to be cases of errors or artifacts in the data on drug quantities, which manifest as an implausible spike in prescribing from a particular trust such that it makes up 90% of all prescribing nationally for a given month. I hope that making these effects more accessible, these issues may be made more visible and thereby corrected or avoided.

## Future plans

My plans for extending this tool are limited. I agree with the sentiment (Goldacre, 2023) that core web services such as this should be developed and delivered by teams including professional software engineers and people with specialised domain knowledge. I hope that a similar service will soon be run by such a team, for example at Open Prescribing (Open Prescribing Hospitals). In the meantime I welcome pull requests, but am limited in my resources for extending the application. I plan to regularly update the dataset with future SCMD releases.

I hope that my tool can be useful, both in providing an opportunity to analyse this data until other similar tools exist, and in some of the features it provides, such as *Ingredients* mode, which I believe does not have a direct analog in OpenPrescribing, apart from in specific measures.

The code behind this tool is openly available at https://github.com/theosanderson/ hospitalprescriptions/ and has an MIT License.

## Data Availability

Data is obtained from a public source: SCMD

https://hospitalprescriptions.genomium.org/

https://opendata.nhsbsa.net/dataset/secondary-care-medicines-data-indicative-price

## Acknowledgements

I’m grateful to the various people who have tried the early version of this tool and provided feedback, including some who pulled out examples I have used in this piece, including: Michael Absoud, Tom Ellis, Luke Jelen, Luke O’Shea and Peter Sivey. I also thank everyone involved in compiling the data that this tool provides access to. This preprint uses a template by Steve Royle and Ricardo Henriques. ChatGPT may have been used for coding, and brainstorming of writing and analyses.

